# Malocclusion Following Early Primary Tooth Extraction: The Role of Socio-Economic Factors and Parental Awareness in Bangladesh

**DOI:** 10.1101/2025.02.27.25323050

**Authors:** Nahian Hossain

## Abstract

The research explored the link between premature deciduous teeth removal and malocclusion as well as parent decision-making affected by socioeconomic factors and their knowledge about early primary tooth extraction and its consequences. A study with 308 child-parent pairs evaluated both urban and rural areas of Bangladesh. Clinical examinations of the children were conducted while parents answered structured questionnaires about the matter. Descriptive statistics is conducted along with Chi-Square tests and logistic regression analyses for data examination. Research findings confirmed that early deciduous tooth extraction caused an increased risk of developing malocclusion since 50.0% of patients in the extraction group developed it compared to 26.0% in the non-extraction group (x^2^ = 6.519, p = 0.011). Parental decision to invest in orthodontic care was influenced by both family financial status and living in an urban area. Higher household earnings (OR = 1.69, p < 0.001) and residing in a city (OR = 7.17, p < 0.001) were discovered as main predictors for willingness to invest. The analysis revealed that 80.5% of parents remained unaware about the connection between early tooth extraction and malocclusion but higher education levels (OR = 0.744, p < 0.001) and urban residence (OR = 0.372, p = 0.005) increased their probability to have this knowledge. Protecting primary teeth prevents malocclusion and demands strategic education programs for parents primarily in rural areas where population has limited education exposure. This study also illustrates how socio-economic factors affect oral health results, which indicates the importance for governments to address limited affordable orthodontic care availability among vulnerable groups in Bangladesh.

## Introduction

Nearly half of the world’s population is affected by oral disease, with most cases occurring in low and middle income countries like Bangladesh (*WHO Highlights Oral Health Neglect Affecting Nearly Half of the World’s Population*, n.d.). Primary teeth are absolutely essential for maintaining proper oral functionality and guiding permanent teeth into proper alignment. Early extraction of primary teeth may lead to malocclusion (Pedersen et al., 1978), “Malocclusion is defined as any deviation from the normal alignment of teeth and the way the upper and lower teeth fit together (occlusion). It includes a range of dental irregularities, such as crowding, spacing, overbite, under bite, cross bite, and open bite, which can affect function, aesthetics, and overall oral health”(Proffit et al., 2019).

Parents must understand the importance of primary tooth retention to prevent early tooth extraction from occurring. Previous research shows that parental oral health knowledge directly affects both child dental health outcomes and their maintained dental care practices (Kaushik & sood, n.d.).

An insufficient knowledge among parents about basic dental care practices, as well as delayed dentist visits and the prolonged effects of untreated dental caries, leads to dental health issues (Kumthekar et al., 2024; Oredugba et al., 2014). Socio-economic factors, such as educational level, income, and residential location, play a significant role in the choices parents make concerning the oral health of their children (Fadila et al., 2024). In Bangladesh, there is a huge gap in healthcare access and awareness. The topic of premature tooth extraction and its effects on malocclusion has not been thoroughly investigated. While some international research addresses the impact of socio-economic and educational determinants on oral health outcomes (Tellez et al., 2014), there is a huge lack of localized data.

It is important to fill this gap for the development of targeted interventions that prioritize both preventive measures and educational initiatives.

The goal of this study is to explore how parental awareness and socio-economic status influence the occurrence of malocclusion resulting from early tooth extraction in children. By examining the relationship between these variables, this research intends to offer data-driven recommendations for policymakers and healthcare professionals aimed at enhancing oral health outcomes in Bangladesh.

### Research Objectives

1. To evaluate the relationship between early tooth extraction and the prevalence of malocclusion in children in Bangladesh.
2. Analyze the socio-economic factors (income, education, residence) that influence parental decisions regarding orthodontic treatment.
3. To investigate the level of understanding among parents regarding the association between early deciduous tooth extraction and the development of malocclusion, and to identify the socio- economic factors (education, residence) that influence parental awareness.

## Methodology

### Study Design

Cross-sectional data collection served this study to evaluate the link between deciduous teeth extraction before age six and malocclusion development as well as parental financing factors and their awareness of extraction outcomes.

### Study Population and Sampling

The research included 308 pairs of children and parents between 6 and 12 years of age from Urban and rural regions of Bangladesh. A random sampling technique was deployed to select Participants, resulting in a representative sample of the target population. Sample size was Determined based on practical constraints and analysis requirements to achieve sufficient Statistical power

### Data Collection

#### Clinical Examination

Trained dental professionals conducted clinical examinations of the children to assess the presence or absence of malocclusion and inputted those data in clinical Data Collection Form. Child’s Gender: (Male /Female), 2) Does the child have malocclusion? (Yes/ No)

#### Survey Questionnaire

A pilot test was conducted with 20 parents to ensure the clarity and validity of the survey instrument. A structured questionnaire was administered to the parents to collect data using following questions: 1) what is your gender? (Male/Female), 2) what is your age? 3) Where do you live? (Urban/Rural), 4) How many children do you have?, 5) What is the highest level of education you have completed?(No formal education/ Can read and write/ Primary school/ Secondary school/ Higher secondary school/ Bachelor’s degree/ Master’s degree or higher), 6) What is your monthly household income?(Below 5,000 BDT/ 5,000–20,000 BDT/ 20,000–35,000 BDT/ 35,000–50,000 BDT/ Above 50,000 BDT), 7) Do either of the child’s parents have a history of malocclusion?(Yes/No), 8) Has your child ever had a baby tooth (deciduous tooth) removed by a dentist, parent, or anyone else before it fell out naturally?(Yes/No), 9) At what age was the tooth extraction done? (Below 4 years/ 4–6 years/ 7–10 years /11+ years/ N/A), 10) What was the primary reason for the extraction?(Tooth decay/ Trauma or injury/ Tooth was loose and removed at home/ Delayed exfoliation/ Dentist recommended for alignment/ Cultural belief /N/A), 11) Who made the decision to extract the tooth?(Dentist / Local/non-professional practitioner/ Parents/ Child/ N/A), 12) After extraction, did a dentist recommend a space maintainer? (Yes and followed/ Yes but did not follow/ No/ Not aware/ N/A), 13) Do you know early tooth extraction can lead to malocclusion? (Yes/No), 14) Have you discussed space maintenance with a dentist? (Yes/ No/ N/A), 15) Does your child have habits affecting tooth alignment? (Thumb-sucking/ Prolonged pacifier use/ Tongue thrusting/ Mouth breathing/ Nail-biting/ Lip sucking/biting/ Cheek biting/ No Habit), 16) would you invest in orthodontic treatment if recommended? (Yes/ No)

### Variables

#### Dependent Variables

Presence of malocclusion (Yes/No).

Parental awareness of the consequences of early tooth extraction (Yes/No). Parental decisions regarding orthodontic treatment (Yes/No).

#### Independent Variables

Early deciduous tooth extraction (Yes/No).

#### Socio-economic factors

Income (below 5,000 BDT, 5000–20000 BDT, 20000–35000 BDT, 35000–50000 BDT, above 50000 BDT).

Education level (no formal education, can read and write, primary school, secondary school, higher secondary school, bachelor’s degree, master’s degree or higher).

Residence (urban/rural).

Parental history of malocclusion (Yes/No).

Children’s oral habits affecting tooth alignment (thumb sucking/mouth breathing/tongue thrusting/nail biting/lip sucking/prolonged pacifier use/cheek biting).

### Data Analysis

Data were analyzed using SPSS version 26.0. The following statistical methods were employed.

#### Descriptive Statistics

Frequencies and percentages were calculated for categorical variables “such as: malocclusion prevalence, parental awareness, and socio-economic factors”.

#### Inferential Statistics

Previous study used the Chi-Square Test to analyze the relationship between premature loss of deciduous teeth and occlusal relationships in children(Martins-Júnior et al., 2017). Similar approach is taken in this research. Chi-Square Test is used to examine the association between early deciduous tooth extraction and malocclusion. Logistic Regression is used to analyze the influence of socio-economic factors (education, income, residence) on parental decisions regarding orthodontic treatment and the influence of socio-economic factors (education, residence) on parental awareness of the consequences of early tooth extraction.

#### Exclusion Criteria

Cases where parents did not have malocclusion, children had no oral habits, or children underwent early tooth extraction without developing malocclusion were excluded to isolate the relationship between early tooth loss and malocclusion.

## Results

### Descriptive Statistics (Frequencies)

An assessment was conducted with 308 parent-child pairs. Of the 308 children participating, 53.2% were male and 46.8% were female shown in Figure 1.

**Figure 1.**
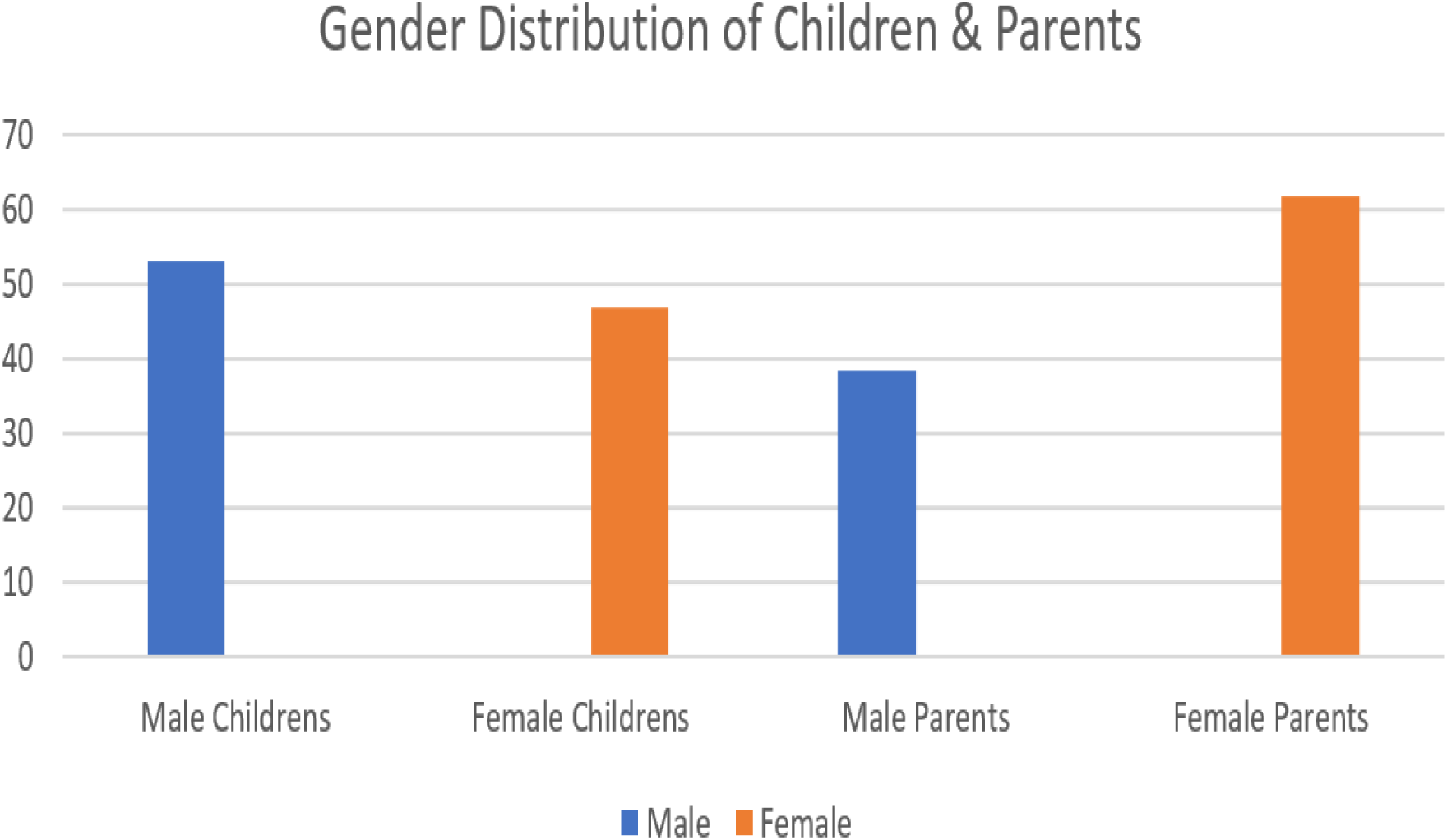
Gender Distribution of Child and Parents

Out of the participating parents who completed the survey a majority identified as female at 61.7% (Figure 1) while 54.9% of families lived in rural areas. The educational levels of parents demonstrated the highest numbers of individuals who finished their education at higher secondary school (27.6%) or secondary school (24.0%) as shown in Figure 2.

**Figure 2.**
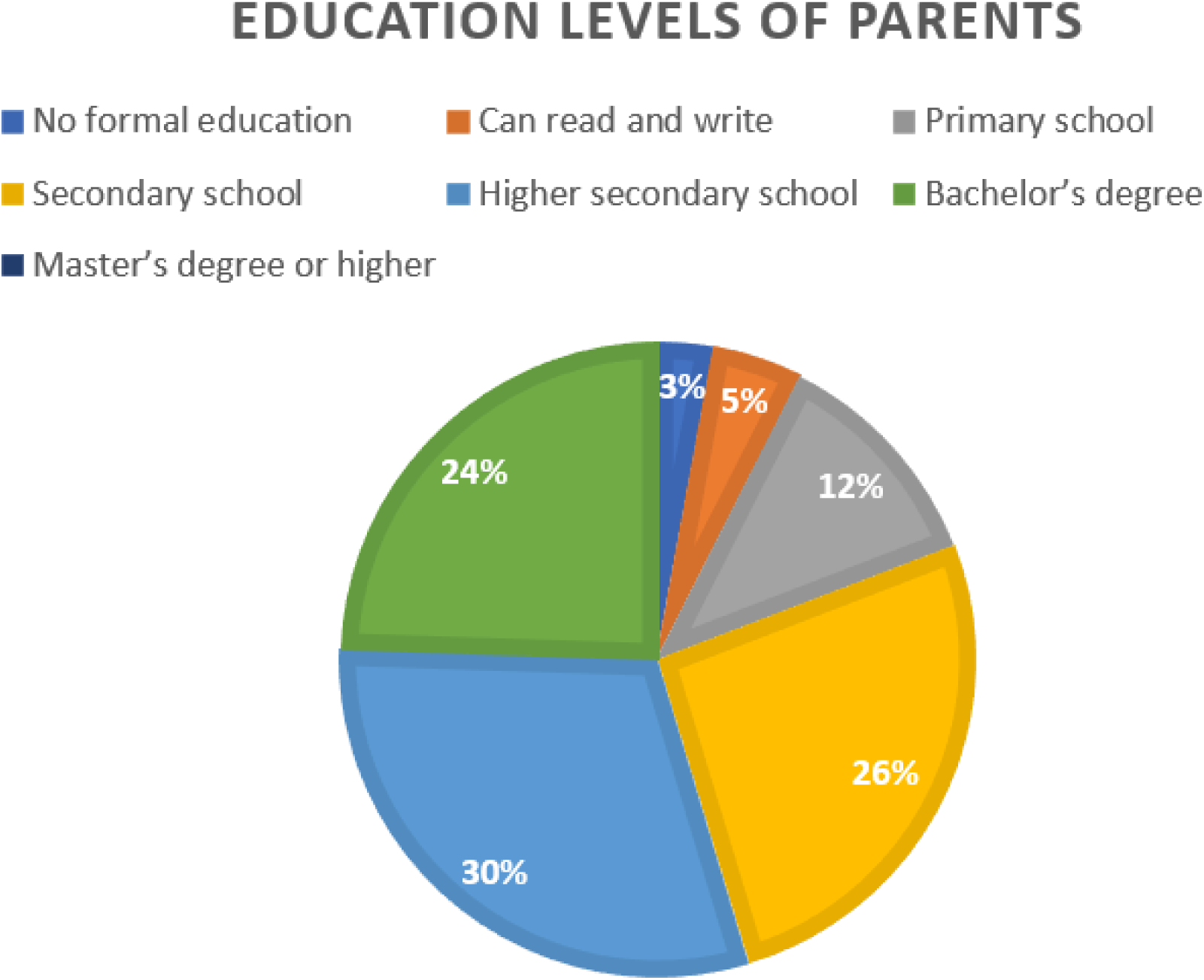

The biggest demographic earned more than 50,000 BDT each month while representing 31.8% of all respondents. 33.8% of parents noticed a past dental irregularity issue in their children though 66.2% percent of parents did not observe any dental irregularities previously. Among children with habits affecting tooth alignment most (72.7%) did not have any habits yet thumb sucking (5.8%) and mouth breathing (5.5%) were found most frequently in children with habits. -participants while 62.0% received a diagnosis of malocclusion. The study results present a complete description of the research subjects while enabling future researchers to analyze the elements that affect malocclusion. The table provided in (Table 1) and (Table 2) shows the demographic characteristics of study participants and factors related to malocclusion.

**Table 1.**
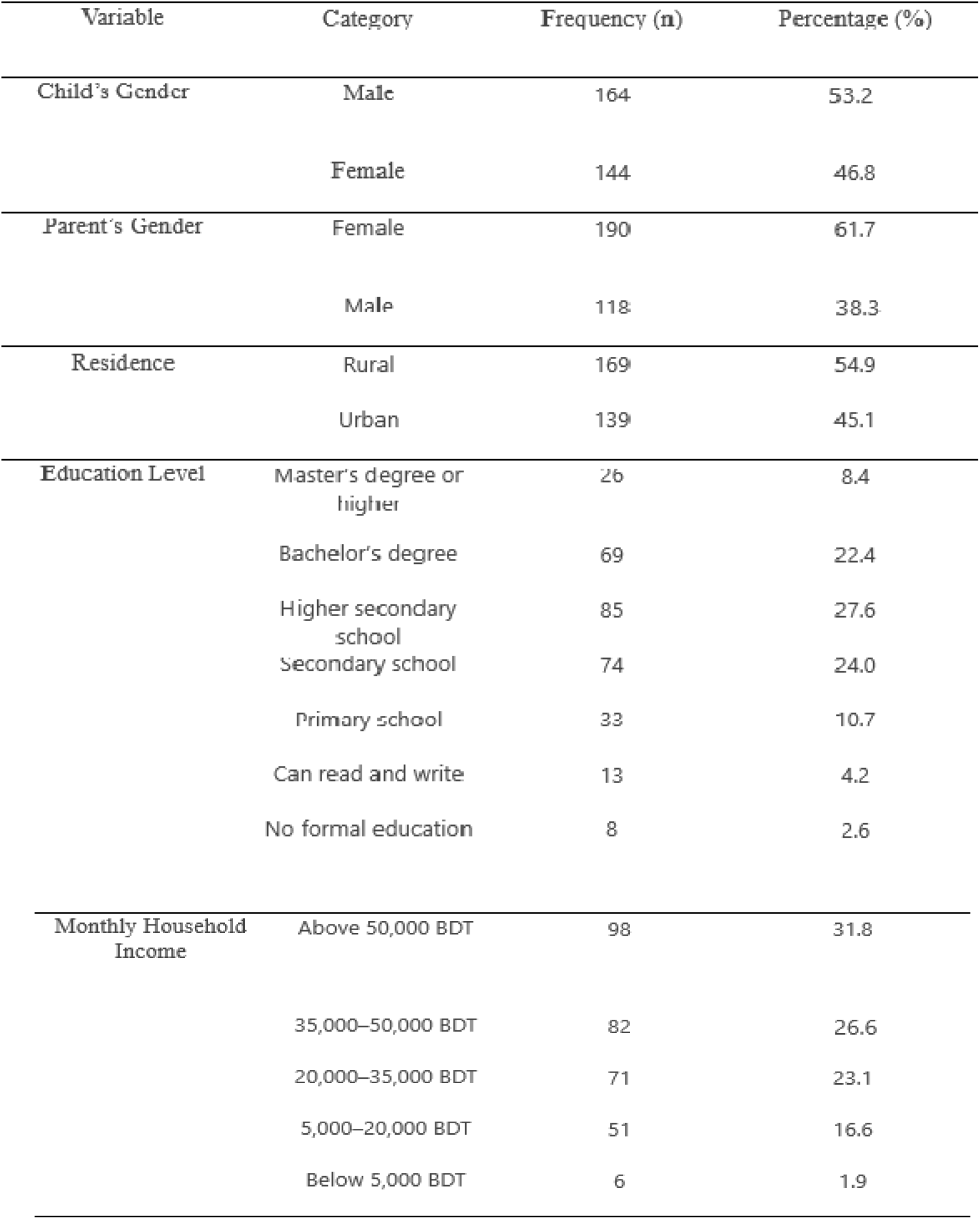
Demographic Characteristics of Study Participants.

**Table 2.** Factors Related to Malocclusion of Study Participants.

| Variable | Category | Frequency (n) | Percentage (%) |
| --- | --- | --- | --- |
| Parental History of Malocclusion | Yes | 104 | 33.8 |
|  | No | 204 | 66.2 |
| Child's Habits Affecting Tooth Alignment | None | 224 | 72.7 |
|  | Thumb sucking | 18 | 5.8 |
|  | Mouth breathing | 17 | 5.5 |
|  | Nail-biting or object-biting | 17 | 5.5 |
|  | Lip sucking or biting | 10 | 3.2 |
|  | Tongue thrusting | 9 | 2.9 |
|  | Prolonged pacifier use | 7 | 2.3 |
|  | Cheek biting | 6 | 1.9 |
| Early Tooth Extraction | Yes | 44 | 14.3 |
|  | No | 264 | 85.7 |
| Child's Malocclusion Status | Yes | 191 | 62.0 |
|  | No | 117 | 38.0 |

### Inferential Statistics

A Chi-Square test was conducted to determine whether early deciduous tooth extraction causes to malocclusion in children. The sample data of 153 children included 123 children who did not have their deciduous teeth removed before they naturally fell out, and 30 who underwent extraction.

The data reveals a statistically important link between early extraction of deciduous teeth and the development of malocclusion (χ² = 6.519, df = 1, p = 0.011), as shown in Table 3. When using the Continuity Correction test the value was 5.440 with p = 0.020 and the Likelihood Ratio test produced χ² = 6.148 while showing p = 0.013. Analysis through Fisher’s Exact Test demonstrated statistical significance using two-tailed p-value 0.015 together with one-tailed p-value 0.011. Results from the Linear-by-Linear Association test confirmed the study findings with a χ² value of 6.476 that achieved statistical significance at p = 0.011.

**Table 3:** Chi-Square Test Results for Early Tooth Extraction and Malocclusion.

| Chi-Square Tests | Value | df | Asymptotic Sig. (2-sided) | Exact Sig. (2-sided) | Exact Sig. (1-sided) |
| --- | --- | --- | --- | --- | --- |
| Pearson Chi-Square | 6.519 | 1 | 0.011 | — | — |
| Continuity Correction | 5.440 | 1 | 0.020 | — | — |
| Likelihood Ratio | 6.148 | 1 | 0.013 | — | — |
| Fisher's Exact Test | — | — | — | 0.015 | 0.011 |
| Linear-by-Linear Association | 6.476 | 1 | 0.011 | — | — |
| N of Valid Cases | 153 | — | — | — | — |

The figure provided in Figure 3 shows the prevalence statistics for malocclusion with respect to patients who received tooth extractions during their childhood. The malocclusion development rate among children who received early tooth extractions reached 50.0% while those who did not have early extractions experienced a 26.0% occurrence (Figure3).

**Figure 3.**
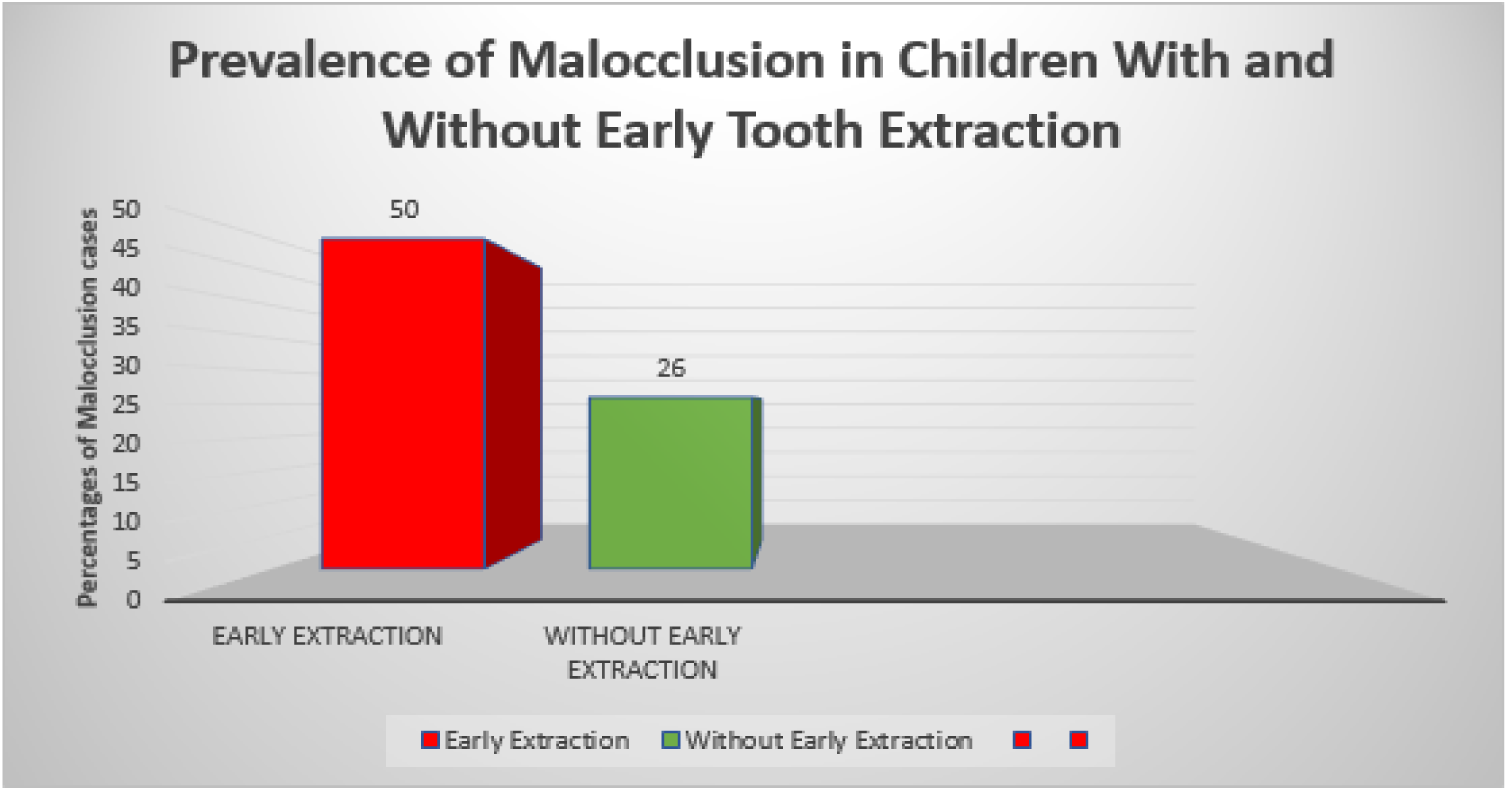
Children who experience early removal of primary teeth tend to develop more cases of malocclusion than children who keep their deciduous teeth. The evidence confirms how premature tooth loss may lead to malocclusion development in children.

### Logistic Regression Analysis

In a previous research which was conducted in China, used logistic regression to examine how education level, income, and residential location influence parental orthodontic treatment choices (Chen et al., 2020). Utilizing similar approach a logistic regression is done to determine how education level together with income and residential location influence parental orthodontic treatment choices. The analysis revealed statistical significance along with 52.8% variance explanation through the χ²(3) = 151.13, p < 0.001 (Nagelkerke R² = 0.528). As shown in Table 4, parents with lower levels of education had less tendency (OR = 0.82, p = 0.009) to get orthodontic treatment compared to parents with higher income levels (OR = 1.69, p < 0.001) and those who resided in urban areas (OR = 7.17, p < 0.001). Parental choices for orthodontic treatment strongly depend on socio-economic characteristics based on the research data.

**Table 4.** Logistic Regression Results for Socioeconomic Factors Influencing Orthodontic Treatment Decisions.

| Variable | B | SE | Wald | df | Sig. | Exp(B) |
| --- | --- | --- | --- | --- | --- | --- |
| Education | -0.204 | 0.078 | 6.739 | 1 | 0.009 | 0.816 |
| Income | 0.523 | 0.106 | 24.142 | 1 | 0.000 | 1.687 |
| Residence<br>(Urban vs.<br>Rural) | 1.970 | 0.330 | 35.632 | 1 | 0.000 | 7.173 |
| Constant | -4.383 | 0.722 | 36.886 | 1 | 0.000 | 0.012 |

### Descriptive Statistics for Parental Awareness

The survey showed that most parents 80.5% did not know how early tooth removal leads to malocclusion as compared to parents who acknowledged this relationship at 19.5% (Table 5).

**Table 5:**
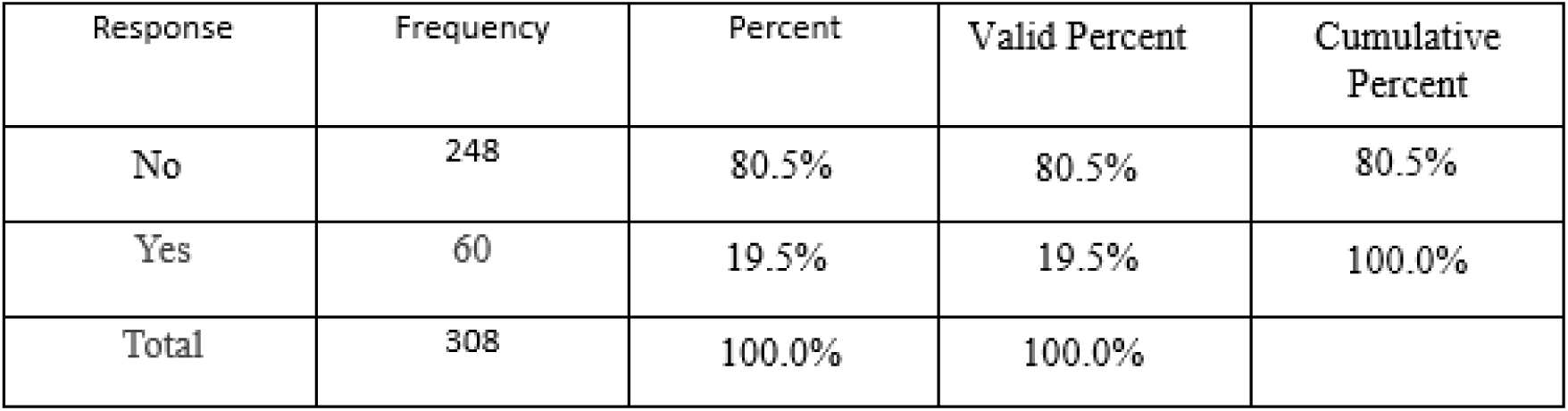
Descriptive Statistics for Parental Awareness of Early Tooth Extraction and Malocclusion.

| Response | Frequency | Percent | Valid Percent | Cumulative Percent |
| --- | --- | --- | --- | --- |
| No | 248 | 80.5% | 80.5% | 80.5% |
| Yes | 60 | 19.5% | 19.5% | 100.0% |
| Total | 308 | 100.0% | 100.0% |  |

Evidence shows that parents possess inadequate awareness about the result from losing early primary teeth and it’s relation to malocclusion.

### Logistic Regression Analysis

A logistic regression analysis evaluated the impact of education combination with residence status as factors which shape parental understanding regarding the relationship between tooth removal and malocclusion during early ages. Results show the statistical significance of this model with χ² = 38.057 (df = 2) p < 0.001 determining parental awareness through 11.6% to 18.5% of explained variance (Cox & Snell R² = 0.116, Nagelkerke R² = 0.185). As shown in Table 6 the research findings proved that education status and place of living acted as vital factors that influenced parental knowledge levels. Parents who completed their education demonstrated a decreased probability of 26% to remain unaware about early tooth extraction consequences (OR = 0.744, p < 0.001). Urban parents exhibited less absence of awareness than rural parents (OR = 0.372, p = 0.005) by 62.8%.

**Table 6.** Combined influence of multiple socio-economic factors (e.g., education, residence) on parental awareness.

| Variable | B | SE | Wald | df | Sig. | Exp(B)<br>(Odds<br>Ratio) |
| --- | --- | --- | --- | --- | --- | --- |
| Residence<br>(Urban vs.<br>Rural) | -0.988 | 0.348 | 8.052 | 1 | 0.005 | 0.372 |
| Education | -0.296 | 0.085 | 12.124 | 1 | 0.000 | 0.744 |
| Constant | 0.006 | 0.277 | 0.001 | 1 | 0.982 | 1.006 |

### Discussion

This study aimed to find out the relationship between early deciduous tooth extraction and malocclusion, the influence of socio-economic factors on parental decisions regarding orthodontic treatment, and the level of parental awareness about the consequences of early tooth extraction. The findings provide valuable insights into the factors contributing to malocclusion in children and the socio-economic barriers to oral health care in Bangladesh.

## Key Findings and Interpretation

Research demonstrated a strong link between premature extraction of primary teeth and malocclusion formation as a result. Children who went through early primary tooth extraction developed malocclusion in 50.0% of cases while children without early primary tooth extraction only showed malocclusion in 26.0% of cases. The results demonstrate the importance of primary teeth because they help direct permanent teeth into proper alignment (Nanda & Kapila, 2010).

Preventive dental measures become essential to keep primary teeth because extraction cases reveal higher malocclusion rates if space maintainer is not used, which demonstrate that tooth preservation should remain a priority and premature tooth loss needs to be avoided(Tabatabai & Kjellberg, 2023).

Recent research demonstrates that socioeconomic elements as well as income levels and basic education along with residential district affect how parents decide about orthodontic care for their children (Almarhoumi, 2024). People earning more money in addition to living in urban areas showed increased inclination toward getting orthodontic treatment yet people with little formal education showed decreased inclination. Research evidence shows that financial obstacles along with restricted healthcare provisions create obstacles for people to receive oral healthcare (Almarhoumi, 2024; Vujicic et al., 2016). Parents living in urban areas usually had superior dental service accessibility along with higher income levels so they frequently chose to get their child orthodontic treatment. Parents in rural areas face more challenges because they lacked adequate dental services combined and financial barriers.

A substantial lack of awareness existed among parents about what happens after their children get early tooth extraction. Research revealed that 80.5% of participating parents failed to understand that infantile tooth extraction leads to malocclusion development but 19.5% correctly recognized this link. Parents residing in urban areas and attaining higher education demonstrated better understanding of long-term effects from early tooth loss based on research conducted in Ludhiana. Research data demonstrates that educational programs should focus immediately on increasing parental awareness about tooth extraction effects since rural families have minimal understanding of these consequences.

### Implications for Policy and Practice

The research outcomes have substantial implications which affect how government supports oral health services alongside persistent dental practices in Bangladesh. The research demonstrates that, early tooth extraction strongly relates to the development of malocclusion (Pedersen et al., 1978)[2], which highlights the importance of preventing dental extraction of primary teeth or preserving primary teeth. Dentist and healthcare providers need to emphasize primary tooth significance during patient evaluations while teaching parents about loss of teeth’s adverse effects (Law, 2013). The public health sector must launch initiatives which teach people about how primary teeth affect both the correct positioning of permanent teeth and their proper alignment (López-Gómez et al., 2016). The socio- economic conditions of families require complete orthodontic access with financial assistance programs especially for underserved low-income families residing in rural areas. Underneath the current framework policymakers should develop specific benefits or insurance systems to lower orthodontic treatment costs for less privileged population segments. The expansion of dental services in rural locations would help equally distribute dental care to urban and rural communities.

Educational programs must be developed to raise parental understanding about how early tooth extraction affects their children. Strategic platforms including educational institutions together with community and healthcare facilities could deliver information about primary tooth importance alongside the possible hazards from premature teeth removal. Material about dental care awareness including brochures and videos needs to be distributed to parents mainly in rural areas since these regions show the lowest understanding about dental consequences.

## Limitations

This study delivers important knowledge yet it contains certain boundary restrictions. The use of a cross-sectional design prevents us from demonstrating that early tooth extraction leads to malocclusion development because it lacks causal relationship identification abilities. Follow-up research through longitudinal studies needs to validate the current analysis while measuring the persistent effects of initial tooth extraction. Self-reported data about parental awareness and socioeconomic information introduces recall bias to the study since researchers depend on these subjective measurements. Future research should adopt objective verification methods such as dental records together with income verification systems in order to minimize biased reporting. Generalization of research results posed a limitation because the data collection occurred within a specific region of Bangladesh.

## Future Research Directions

Future studies should investigate other elements which affect parental decision-making processes from a cultural standpoint and their ability to obtain dental services and provider involvement in dental education. Studies following the same participants from birth would show the exact relationship that exists between the early extraction of teeth and malocclusion’s development.

Research should examine how well various interventions increase both parent understanding of dental care and their ability to obtain orthodontic treatments for children. Research conducted in diverse regions of Bangladesh would expose any differences between districts while helping develop interventions for specific areas.

## Conclusion

This research investigated the malocclusion linked to premature primary tooth removal together with social status effects on orthodontic selections by caregivers and their comprehension of dental extraction outcomes. Early tooth extraction demonstrated a direct link to malocclusion (Pedersen et al., 1978) since 50.0% of extracted children developed malocclusion but the non-extraction group had 26.0%. The parental choice for orthodontic treatment is determined by both socio-economic variables like income level and education status and residential area since urban parents who earned higher incomes frequently choose orthodontic treatment. The research showed parents remain uninformed about the risk of malocclusion after early tooth extraction because 80.5% of parents were ignorant about this connection. These research findings have significant implication for the development of policies and practices regarding oral health throughout Bangladesh. Preventive dental care is important because early extraction of teeth demonstrates its strong relationship with malocclusion issues. The public healthcare system needs to educate rural and uneducated parents regarding primary teeth value and their exposed dangers following premature extractions. The government needs to establish programs which enhance affordability of orthodontic treatments for families from rural areas and those who earn low income.

Further research into parental decision-making and awareness is needed, because this research did not examined cultural beliefs and dental care access. Research over extended periods must verify how early tooth extraction affects malocclusion development and test methods to enhance oral health results.

In conclusion, this research contributes to developing body of evidence regarding primary teeth importance together with socio-economic oral health barriers in developing nations.

Policymakers together with healthcare providers can decrease malocclusion prevalence and enhance child oral health results through barrier elimination and parental education in Bangladesh.

## Data Availability Statement

The data presented in this study are available on request from the corresponding author. The data are not publicly available due to privacy concerns.

## Ethical approval and consent to participate

In accordance with ethical considerations and regulatory guidelines, the research ethics committee of the Bangladesh Medical Research Council (BMRC) has granted approval for the study protocol under reference number (774-06-24, approval date 14/06/2024). Additionally, prior to participation in the study, informed consent was obtained from all parents or legal guardians of the participants. This ensured that participants’ rights and well-being were safeguarded throughout the research process. While the study is an observational cross-sectional design and does not involve experiments on humans or the use of human tissue samples, we have taken proactive measures to uphold ethical standards and maintain transparency in our methodology.

## Conflict of interest

The authors declare no conflict of interest.

## Funding Disclosure

This research received no external funding

## Notes

### Competing Interest Statement

The authors have declared no competing interest.

### Funding Statement

No external funding was received

### Author Declarations

Ethics Committee of the Bangladesh Medical Research Council (BMRC) gave ethical approval for this work.

### Summary of Updates

Corrected factual error in the sentence 'protecting permanent teeth prevents malocclusion' to 'protecting primary teeth prevents malocclusion' Corrected formatting of the Chi-square symbol from χX square to proper notation of x2

